# ADIPOSE-DERIVED STROMAL VASCULAR FRACTION AND FAT GRAFTS VERSUS MEDICAL TREATMENT FOR TREATING THE HANDS OF PATIENTS WITH SYSTEMIC SCLEROSIS. A RANDOMIZED CRONTOLLED TRIAL

**DOI:** 10.1101/2021.02.08.21250465

**Authors:** Martin Iglesias, Iván Torre-Villalvazo, Patricia Butrón-Gandarillas, Tatiana S. Rodríguez-Reyna, Erik A. Torre-Anaya, Armando R. Tovar-Palacio, Alejandro Zentella-Dehesa, Martha Guevara-Cruz, Alan M Hérnandez-Campos

**Affiliations:** Plastic and Reconstructive Surgery Service at Instituto Nacional de Ciencias Médicas y Nutrición, Salvador Zubirán; and Tecnologico de Monterrey, Escuela de Medicina y Ciencias de la Salud, Mexico City, Mexico; Nutrition Physiology Department at Instituto Nacional de Ciencias Médicas y Nutrición Salvador Zubirán, Mexico City, Mexico; Plastic and Reconstructive Surgery Service at Instituto Nacional de Ciencias Médicas y Nutrición Salvador Zubirán, Mexico City, Mexico; Rheumatology Department at Instituto Nacional de Ciencias Médicas y Nutrición Salvador Zubirán, Mexico City, Mexico; Biochemistry Laboratory at Instituto Nacional de Ciencias Médicas y Nutrición Salvador Zubirán, Mexico City, Mexico

**Keywords:** hand therapy, adipose derived vascular stromal fraction, systemic sclerosis, adipose mesenchymal stem cells, regenerative medicine

## Abstract

**Background:** In the hand, the Systemic Sclerosis (SS) is characteristically evidenced by Raynaud’s phenomenon (RP) and fibrosis of the skin, tendons, ligaments, and joints as well as digital ulcers with prolonged healing. Current medical treatment not always cure these complications. Local adipose-derived stromal vascular fraction (ADSVF) administration into the hands has been proposed as an emerging treatment for these complications, due to its proangiogenic, antifibrotic, and immunoregulatory activities. The objective of this controlled trial was to evaluate the safety and clinical effects of fat micrografts plus ADSVF administration into the hands of patients with SS.

**Methods:** This was an open-label, monocentric, randomized controlled study. Twenty patients diagnosed with SS were enrolled and assigned to the experimental or control group. Fat micrografts plus the ADSVF were injected into the right hand of experimental group patients. The control group continued to receive only medical treatment. Demographic, serologic data and disease severity were recorded. Digital oximetry, pain, Raynaud phenomenon (RP), digital ulcer healing (DUH), mobility, thumb opposition, vascular density of the nail bed, skin affection of the hand, Serologic antibodies, hand function, and quality of life scores were evaluated in both groups. The mean follow-up period was 168 days.

The differences between before and after the intervention were analyzed with the Wilcoxon range test, and the differences between the control and experimental groups at 0 days and 168 days were analyzed with the Mann–Whitney U test.

**Results:** Adverse events were not observed in both groups. There were no changes in disease severity, serologic antibodies, nailfold capillaroscopy patterns, mobility, and hand function in both groups. There were significant improvements in pain, DUH and quality of life scores in the experimental group. RP improved significantly in both groups. However, on statistically comparing the results at 168 days between the groups, significant improvements were only observed in pain levels (*p* = 0.02) and DUH (*p*=0.003).

**Conclusions:** The injection of ADSVF plus fat micrografts is a reproducible, and safe technique. Pain and digital ulcers in the hands of patients with SS can be treated with this treatment.

**Trial Registration:** Retrospectively registered in ClinicalTrials.gov with identifier NCT04387825

## INTRODUCTION

Systemic sclerosis (SS) is characterized by the presence of functional and structural microvasculopathy resulting in ischemia that leads to cutaneous and visceral fibrosis. In the hand, this disease is characteristically evidenced by Raynaud’s phenomenon (RP) and fibrosis of the skin, tendons, ligaments, and joints.[1,2,3,4] Further, the hand is affected in 47 to 97% of the SS cases.[5, 6] Permanent ischemia and skin fibrosis can lead to the appearance of digital ulcers that result in prolonged healing durations, recurrences, and infections and may culminate in digital amputation. This musculoskeletal involvement is one of the main etiologies of pain, devastating disability, and a dramatic decrease in the quality of life of patients with SS.[6, 7] The current treatment is the control of the disease with medications and rehabilitation.[8] However, these options only temporarily control the symptoms but do not decrease the time period required for ulcer healing or improve hand function.

The presence of adipose stem cells (ASCs) in the adipose-derived stromal vascular fraction (ADSVF) was described by Zuk et al. in 2002.[9] Because these cells are multipotent with local angiogenic, anti-inflammatory, antifibrotic, immunomodulatory, and regenerative properties and because of their abundance in fat, easy acquisition, and almost immediate availability for use, ADSVF administration has positioned itself as an alternative treatment for immunological diseases that cause ischemia and cutaneous fibrosis, which are also the main manifestations of SS.[9-21]

Bank et al. infiltrated decanted fat only in order to treat RP and obtained extraordinary results in terms of improving pain and RP.[22] Del Papa et al. centrifuged fat and injected the intermediate layer into patients with SS and active digital ulcers that were resistant to conventional treatment.[14, 15] Subsequently, Del Papa et al. reported on the same technique in a controlled study in 2019. The digital ulcers healed after a mean period of 8 weeks with the reduction of pain and formation of new capillaries.[23]

Granel et al. administered the ADSVF to 12 patients who predominantly had limited cutaneous SS with no severe involvement of the internal organs. Subsequently, they reported improvements in pain and the frequency and intensity of RP; the healing of digital ulcers that were resistant to conventional treatments; improved strength, movement, and Cochin functional scale scores; and a decrease in the digital circumference.[24] Further, these results were reported to persist for 12, 22, and 30 months postoperatively with additional improvements in nail appearance and health based on improved Scleroderma Health Assessment Questionnaire scores, the ability to participate in household activities, and the approval of the procedure by the patients themselves.[21, 25, 26] Del Papa et al. and Granel et al. reported that the treatment was safe and well tolerated.[14, 24]

Lipoatrophy may develop in patients with SS due to the loss of adipocytes and defective stem cell function.[13, 27] Thus, there is a good possibility that the hands of patients with SS could be treated with ADSVF plus fat grafts with the from proximal sites, such as the abdominal wall. For these reasons and because there has been only one controlled trial, we designed this controlled clinical trial with the aim of evaluating the safety, reproducibility, and clinical effects of treating the hands of patients affected with SS with the administration ADSVF plus fat micrografts.

## METHODS

This open-label, monocentric, randomized controlled pilot study was approved by the Institutional Review Committee with the reference, SCI-1505-15/15-1, and was registered with ClinicalTrials.gov under the identifier, NCT04387825. We used a random number table to assign patients into different groups. Informed consent to participate in this study was obtained from all the paticipants. Both the experimental and control groups contained 10 patients each who were diagnosed with SS according to the criteria of the American College of Rheumatology and LeRoy-Medsger criteria. All the patients were aged > 18 years and had BMIs > 18 kg/m^2^. All the patients had undergone stable vasoactive and immunosuppressive therapies for at least 1 month before enrollment in the study, and these treatments were continued unchanged throughout the study period. The exclusion criteria were infected digital ulcers, comorbidities that could affect hand function, alcoholism, and/or drug abuse. It was decided that the treatment would be administered to the most affected hand of the patients, which was the right hand in the entire experimental group. ADSVF and fat micrografts were administered in the experimental group. The evolution of the disease and the medical therapy effects were observed in the control group. Demographic and serologic data were recorded as well as disease severity according to the Medsger severity scale. The evaluated variables and follow-up times are presented in Table 1.

**Table 1.** Data Evaluation.

| Variable | Evaluation method | Follow up in days |  |  |  |  |  |  |
| --- | --- | --- | --- | --- | --- | --- | --- | --- |
|  |  | Preop | 28 | 56 | 84 | 112 | 140 | 168 |
| Digital Oximetry | Transcutaneous Oximetry | ✓ | ✓ | ✓ | ✓ | ✓ | ✓ | ✓ |
| Pain | Numeric scale from 1 to 10 | ✓ | ✓ | ✓ | ✓ | ✓ | ✓ | ✓ |
| Raynaud Phenomenon | Frequency number of events per day/week; Duration of minutes in every event. | ✓ | ✓ | ✓ | ✓ | ✓ | ✓ | ✓ |
| Digital Ulcers | Number of ulcers | ✓ | ✓ | ✓ | ✓ | ✓ | ✓ | ✓ |
| Digital TAM* | Digital Manual Goniometry | ✓ |  |  |  |  |  | ✓ |
| Thumb opposition | Kapandji Test | ✓ |  |  |  |  |  | ✓ |
| Health status and disability index | SHAQ scale | ✓ |  |  |  |  |  | ✓ |
| Hand Function | COCHIN scale | ✓ |  |  |  |  |  | ✓ |
| Quality of Life | SF-36 scale | ✓ |  |  |  |  |  | ✓ |
| Vascular density of the nail bed | Nailfold Videocapillaroscopy | ✓ |  |  |  |  |  | ✓ |
| Skin affection of the hand | Modified Rodnan skin score | ✓ |  |  |  |  |  | ✓ |
| Antibodies | ANA**, Anti-Topoisomerase I, Anti-centromere, Anti-U1-RNP | ✓ |  |  |  |  |  | ✓ |

\*TAM=Total Active Motion
\*\*ANA=Anti-nuclear antibodies

Data regarding the presence of complications, such as compartment syndrome and/or infection, in both the donor site and treated hand were recorded daily for 1 week.

Rheumatological data were retrieved by the rheumatologist group, and medical interns were trained to allocated patients and collect the other data.

### Tissue collection and ADSVF preparation

Fat grafts were obtained by liposuction under local anesthesia. Briefly, the periumbilical region (9 patients) or posterior thigh (1 patient) was infiltrated with 500 ml of Klein’s formula. Liposuction was performed with a 20-ml syringe and 3-mm diameter blunt cannula with 2-mm openings. A total of 100 ml of fat was harvested, from which 60 ml was transferred to sterile flask containing Hank’s balanced saline solution with 5% albumin and was immediately transported to the laboratory. The adipose tissue was decanted for 10 minutes in a laminar flow cabinet for fraction separation. The upper fraction comprised disrupted adipocytes (oil), and the lower fraction was a fluid portion containing the anesthetic solution, erythrocytes, and dead cells. These both fractions were removed. The middle fraction was washed twice in phosphate-buffered saline (PBS) (Gibco, Thermo Fisher Scientific, Waltham, MA, USA) and subsequently incubated for 20 minutes in a type II collagenase solution (Sigma, St. Louis, MO, USA) in PBS at a concentration of 2 mg/ml of lipoaspirate. This was followed by addition of ethylenediaminetetraacetic acid (Sigma) in PBS. To separate the ADSVF from the floating adipocytes, this digested tissue was centrifuged at 800 × *g* for 10 minutes. The compacted cells were resuspended in 3 ml of PBS and filtered through a 100-μm mesh cell strainer (Corning Inc., Corning, NY, USA). A 10 μl aliquot was obtained from each patient to characterize the ADSVF cellular identity by flow cytometric assays. After the fat was processed, 2 ml of the ADSVF was transferred back to the operating room for the autologous transplantation procedure. Another 1 ml sample was used for cell quantification, viability determination, and cell culturing to determine the adipocyte differentiation capacity (Figure 1).

**Figure 1.**
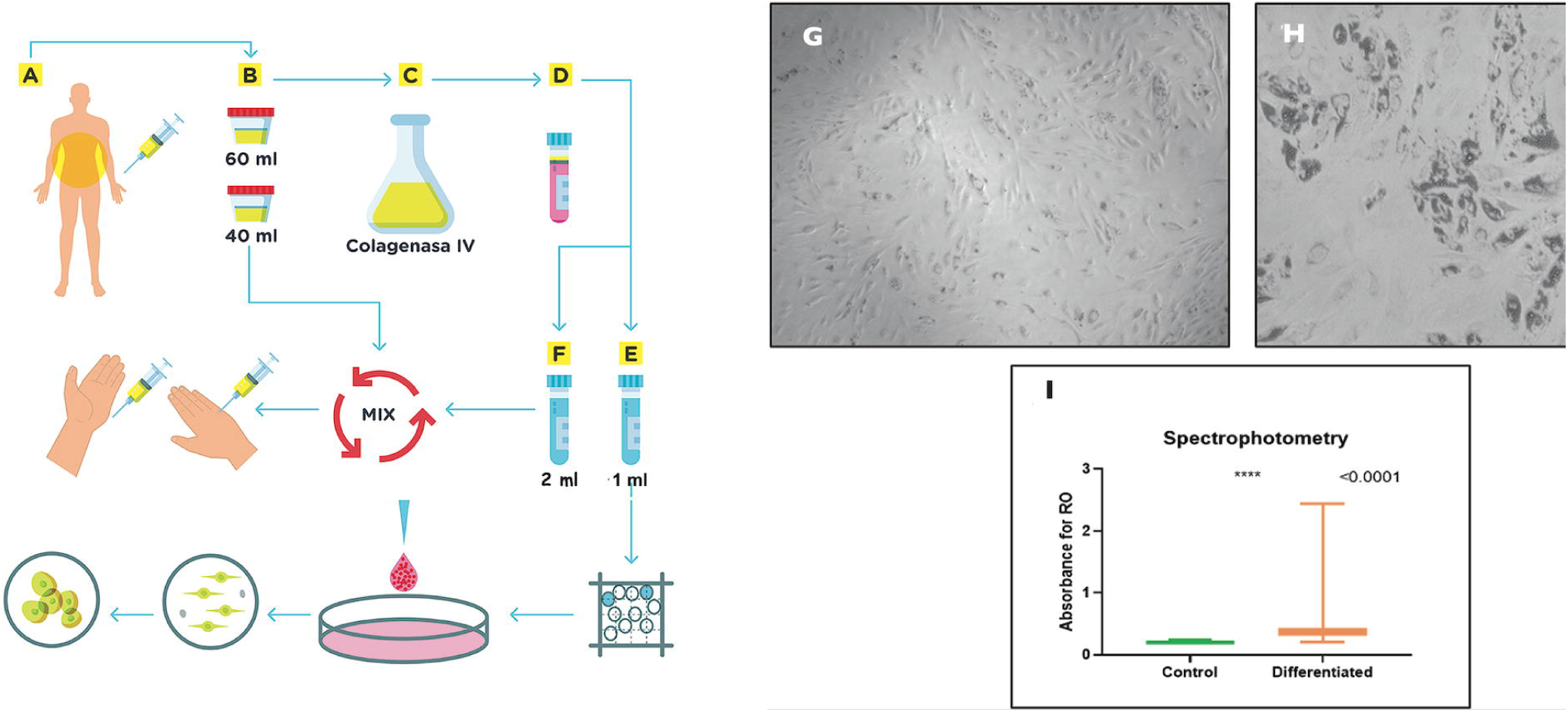
Flow chart of the methodology. (A) Fat grafts are obtained by abdominal liposuction. (B) A 60 ml volume is sent to the laboratory. (C) The enzymatic digestion of fat is performed by a collagenase separation method. (D) The initial adipose-derived stromal vascular fraction (ADSVF) is divided into two samples. (E) The first is a 1 ml sample for the characterization and control of the cell culture. (F) The second is a 2 ml sample that is reserved for administration to the patient in a lipoaspirate mixture by infiltration into the hand. The differentiation process is depicted in the last row. Differentiation. (G) Control culture without adipogenic stimulation stained with oil red (OR) at day 20. (H) Culture with adipogenic stimulation stained with OR on day 20. (I) Spectrophotometric analysis of the two groups depicted in a graphic. There was *a higher OR absorption level in the differentiated group (p* = 0.0001).

The cells were seeded in 6-well culture plates in Dulbecco’s modified Eagle’s medium and were supplemented with fetal bovine serum and antibiotic/antifungal agents. Upon reaching 90% confluence, the cells were differentiated into mature adipocytes using a differentiation cocktail (2.5 µM dexamethasone, 0.5 mM 3-isobutyl-1-methylxanthine, 10 µg/ml insulin). As a control, a plate containing cells in the culture medium was left without adipogenic stimulation. After 20 days of differentiation, the cells were stained with oil red to assess fat accumulation (mature adipocyte phenotype), and the fat content in each culture was quantified by spectrophotometry (Figure 1 A-F).

### ADSVF administration into the hand

A nerve blockade was induced in the median, ulnar, and radial nerves at the wrist level using 2% lidocaine without epinephrine. Subsequently, 40 ml of fat was mixed with 2 ml of the ADSVF and was transferred to 1-ml and 3-ml syringes. Using a 19-gauge blunt cannula (0.8 mm), 1.5 ml was administered along each radial and ulnar neurovascular digital pedicle through small radial and ulnar skin incisions at the level of the interphalangeal and metacarpophalangeal joints. Additionally, 3 ml was administered to each side of the metacarpal trapezius joint, together with the subcutaneous distribution of 10 ml throughout the palm of the hand and even distribution of 10 ml in the back of the hand. The study was carried out from October 2018 to May 2019.

### Statistical analysis

Continuous variables are expressed as median with 95% confidence interval. Dichotomous variables are expressed as frequency and percentage. Categorical or dichotomous variables were analyzed with Fisher’s exact test. The differences between before and after the intervention were analyzed with the Wilcoxon range test, and the differences between the control and experimental groups at 0 days and 168 days were analyzed with the Mann– Whitney U test. The analysis was enhanced to determine the interaction between time (0/168 days) and treatment (control/experimental) with analysis of variance for repeated measures, and logarithmic transformation was performed before such analysis. A P value of <0.05 was considered statistically significant of a tail. The data were analyzed using SPSS for Windows, version 24.00 (IBM Corp., Armonk, NY, USA) and GraphPad Prism software version 7 (GraphPad Software, San Diego, CA, USA).

## RESULTS

Demographic, serological (antibodies), and disease severity-related data are presented in Table 2.

**Table 2.** Demographic, serological and severity data. Control and experimental groups.

| General Data |  | 0 days |  | 168 days |  |  |
| --- | --- | --- | --- | --- | --- | --- |
| Patient (n) | Experimental<br>10 | Control<br>10 | P<sup>1</sup> | Experimental<br>10 | Control<br>9 | P |
| Female n (%) | 10 (100 %) | 9 (95 %) | 0.99 | 10 (100%) | 8 (89%) | 0.47 |
| Age, mean (95%IC)* | 55.0(43.4,58.7) | 57.0(45.6,62.5) | 0.48 | 55.1(46.0,62.7) | 55.0(43.5,59.0) | 0.48 |
| Diffuse Sclerosis, n (%) | 8 (80 %) | 6 (60 %) | 0.62 | 8 (80 %) | 6 (66.66%) | 0.62 |
| Antibodies |  |  |  |  |  |  |
| ANA positives, n (%) | 8 (80 %) | 10 (100%) | 0.47 | 8 (80 %) | 9 (100%) | 0.47 |
| Anti-Topoisomerase I, n (%) | 3 (30 %) | 2 (20%) | 0.99 | 3 (30 %) | 2 (22.2%) | 0.99 |
| Anti-centromere, n (%) | 3 (30 %) | 3 (30%) | 0.99 | 3 (30 %) | 3 (33.3%) | 0.99 |
| Anti-U1-RNP, n (%) | 2 (20 %) | 3 (30%) | 0.99 | 2 (20 %) | 3 (33.3%) | 0.62 |
| Organ involvement |  |  |  |  |  |  |
| Vascular, n (%) | 10 (100 %) | 10 (100 %) | 0.99 | 8 (80 %) | 9 (100%) | 0.47 |
| Severe vascular, n (%) | 6 (60 %) | 7 (70 %) | 0.99 | 2 (20 %) | 4 (44.4%) | 0.35 |
| Articular, n (%) | 10 (100 %) | 10 (100 %) | 0.99 | 10 (100 %) | 9 (100%) | 0.99 |
| Severe articular, n (%) | 6 (60 %) | 2 (20 %) | 0.17 | 3 (30 %) | 3 (33.3%) | 0.99 |
| FTP, mean (95%IC)* | 3.15(2.30,3.71) | 4.00 (2.72,4.39) | 0.21 | 3.50(2.25,4.32) | 3.15(2.81,4.00) | 0.91 |
| Muscular, n (%) | 1 (10 %) | 1 (10%) | 0.99 | 3 (30 %) | 1 (11.1%) | 0.58 |
| Severe muscular, n (%) | 0 (-) | 0 (-) | 0.99 | 0 (-) | 0 (-) | 0.99 |
| Gastrointestinal, n (%) | 4 (40 %) | 5 (50 %) | 0.99 | 5 (50 %) | 7 (77.7%) | 0.35 |
| Severe gastrointestinal, n (%) | 1 (10 %) | 1 (10 %) | 0.99 | 1 (10 %) | 1 (11.1%) | 0.99 |
| Pulmonary, n (%) | 6 (60 %) | 9 (90%) | 0.30 | 6 (60 %) | 9 (100%) | 0.08 |
| Severe pulmonary, n (%) | 1 (10 %) | 1 (10 %) | 0.99 | 1 (10 %) | 1 (11.1%) | 0.99 |
| HAP, n (%) | 2 (20 %) | 3 (30 %) | 0.99 | 2 (20 %) | 3 (33.3%) | 0.62 |
| Severe HAP, n (%) | 0 (-) | 0 (-) | 0.99 | 0 (-) | 0 (-) | 0.99 |
| Cardiac, n (%) | 1 (10 %) | 1 (10 %) | 0.99 | 1 (10 %) | 1 (11.1%) | 0.99 |
| Severe cardiac, n (%) | 0 (-) | 0 (-) | 0.99 | 0 (-) | 0 (-) | 0.99 |
| Renal, n (%) | 0 (-) | 1 (10%) | 0.99 | 0 (-) | 1 (11.1%) | 0.47 |
| Severe renal n (%) | 0 (-) | 0 (-) | 0.99 | 0 (-) | 0 (-) | 0.99 |
| Capillaroscopic Pattern |  |  |  |  |  |  |
| Early phase n(%) | 2 (20 %) | 1 (10 %) | 0.99 | 3 (30 %) | 0 (-) | 0.21 |
| Active phase n(%) | 0 (-) | 1 (10%) | 0.99 | 0 (-) | 1 (11.1%) | 0.47 |
| Late phase n(%) | 8 (80%) | 8 (80 %) | 0.99 | 7 (70 %) | 9 (100%) | 0.21 |

P<sup>1</sup> Statistical significance analyzed Fisher's exact test.
\*Statistical significance analyzed with Mann-Whitney U test

The baseline characteristics were similar between the two patient groups. All the patients presented with moderately intense vascular and joint conditions. Some patients exhibited severe internal organ involvement. One control group patient withdrew from the study due to personal problems.

The total viable nucleated cell numbers and cellular characteristics of the ADSVF are outlined in Additional file 1 Table A.

The abilities to adhere to plastic and differentiate to adipocytes are presented in Figure 1. G-I greater amount of fat content was found in cell cultures induced toward adipocyte differentiation than in control cultures (*p* = 0.0001).

At the completion of ADSVF administration, the digits did not exhibit capillary filling, but circulation was reestablished after 2 h. There were no adverse effects. Digital oximetry (SpO2), digital total active motion, thumb opposition, hand function, health status and disability index, and vascular density of the nail bed, were similar in the two groups at the study’s end.

The pain score increased from 4.6 to 7.5 in the first week, gradually decreased to reach the basal value at day 7, and subsequently began to decrease further. Significant improvements were observed in pain and quality of life score (Short Form 36) in the experimental group at the study’s end. The frequency, intensity and duration of RP have significant improvement in both groups at 168 days, but were more important in the experimental group. The skin affection of the hand improved significantly in the control group. However, when the results at 168 days were statistically compared between the groups, only pain exhibited a significant improvement (p = 0.02) (Table 3 and Additional file 2 Table B).

**Table 3.** Results in control and experimental groups

| Concept | Group | Day 0 | Day 168 | P<sup>1</sup> | P<sup>2</sup> |
| --- | --- | --- | --- | --- | --- |
| Pain | Control | 4.00(2.64,6.16) | 2.00(1.51,6.04) | 0.91 |  |
|  | Experimental | 5.00(3.08,6.12) | 0.00(0.00,2.95)* | **0.006\*** | **0.02\*** |
| Quality of Life<br>(SF-36) | Control | 40.0(28.5,46.4) | 35.0(17.5-51.3) | 0.55 |  |
|  | Experimental | 37.5(32.1-57.8) | 45.0(38.7-62.2) | **0.04\*** | 0.15 |
| Raynaud<br>Phenomenon<br>*Frequency n/week* | Control | 5.50(2.68,13.1) | 00(0.00,03.18) | **0.010\*** |  |
|  | Experimental | 4.50(2.87,6.33) | 0.50(0.10,1.10) | **0.005\*** | 0.418 |
| Raynaud<br>Phenomenon<br>*Intensity* | Control | 2.00(1.22,2.18) | 0.00(0.00,1.44) | **0.03\*** |  |
|  | Experimental | 2.00(1.50,2.10) | 0.50(0.10,1.10) | **0.006\*** | 0.60 |
| Raynaud<br>Phenomenon<br>*duration in*<br>*minutes* | Control | 17.5(2.12,54.4) | 0.00(0.00,10.3) | **0.02\*** |  |
|  | Experimental | 12.5(6.55,35.6) | 0.00(0.00,6.82) | **0.005\*** | 0.14 |
| Skin affection of the<br>hand | Control | 6.50(2.32,15.8)) | 5.50(1.42,12.9) | **0.05\*** |  |
|  | Experimental | 15.0(6.84,17.7) | 15.5(6.36,16.6) | 0.07 | 0.19 |

P<sup>1</sup>= Analyze the differences within groups between baseline and final results. Wilcoxon signed-rank test was used.
\*P<0.05, U de Mann-Whitney analyze the differences between groups.
P<sup>2</sup>= These data were log-transformed before statistical analyses was ANOVA for repeated measures to determine the time x group interaction.

The sample size of the control group n=9, experimental group n=10; Data are presented as median (95% confidence intervals).

The digital ulcers in the experimental group healed within 28 days, while those in the control group healed within 84 days. During the follow-up period, ulcer recurrence was observed in one experimental group patient compared with in three control group patients. These results were statistically significant (Table 4).

**Table 4.** Ischemic Digital Ulcers in Sclerosis Systemic

| Table 4. Ischemic Digital Ulcers in Sclerosis Systemic |  |  |  |  |  |  |  |
| --- | --- | --- | --- | --- | --- | --- | --- |
| Group | Day 0 | Day 28 | Day 56 | Day 84 | Day 112 | Day 140 | Day 168 |
| Experimental (n 10) | 3 | 0 | 0 | 0 | 0 | 1 | 0 |
| Control (n 9) | 4 | 4 | 2 | 1 | 2 | 2 | 6 |
| *p* value* | 0.50 | 0.03 | 0.21 | 0.47 | 0.248 | 0.45 | 0.003 |

\*statistical analysis was performed with the exact fisher test

Clinically, the treated hands of the experimental group patients exhibited larger volumes, warmer temperatures, and faster capillary filling than the contralateral hands of the same patients (Figure 2 and Video 1).

**Figure 2.**
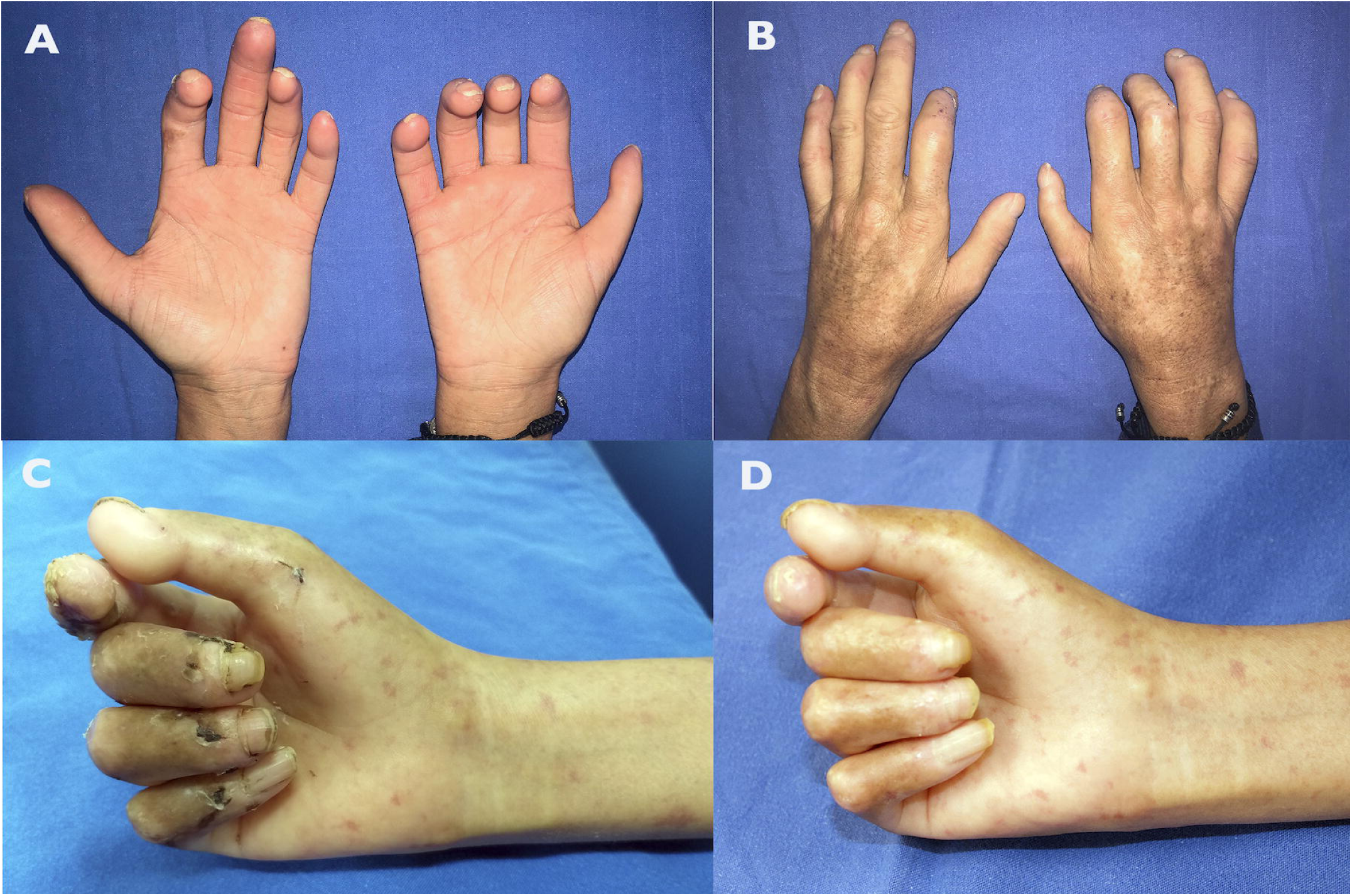
Clinical results. (A, B) Patient 1, female with age range 56-60 y/o. The right hand was injected with micrografts enriched with the adipose-derived stromal vascular fraction. It exhibited a higher volume and was warmer than the left hand. (C) Patient 2, female with age range 31-35 y/o. Clinical status before the treatment. The hand presented with dermo-epidermal abrasions and digital ulcers. (D) The same patient with healing digital ulcers.

Both groups exhibited late capillaroscopic patterns. There was no significant change in the number of nailfold capillary loops between the groups at the study’s end. One experimental group patient exhibited an improved capillaroscopic pattern changing from late to early, while one control group patient exhibited a worsened capillaroscopic pattern changing from early to late.

## DISCUSSION

The application of decanted fat,[22] centrifuged fat,[14] and the ADSVF,[15, 25, 28] has consistently and significantly improved pain, RP, and the healing of digital ulcers in the hands of patients with SS, thereby improving their quality of life. Other inconsistently observed benefits, such as decreased digital circumferences, improvements in digital mobility and strength, improvements in the formation of new subungual capillaries, and improvements in function based on Cochin scale scores, have also been reported.[14, 15, 23]

However, previous studies have been uncontrolled with a short follow-up period of 6 months.[14, 15] There has been only one controlled study that involved the use of decanted grafts.[23] Within 8 weeks, digital ulcer healing was observed in 92% of the experimental group patients compared to in 7.7% of the control group patients. Furthermore, at 20 weeks, pain levels were decreased, and the number of ungual capillaries was increased in the experimental group compared with in the control group.[23]

The present controlled study with a 6-month follow-up period included patients with diffuse and limited SS as well as moderate vascular and articular involvement, including some with severe internal organ involvement. The results observed in the experimental group were similar to those reported in uncontrolled studies. There were statistically significant improvements in pain,, digital ulcer healing and SF-36 scores in the experimental group. However, when the variables were compared with those of the control group, only the improvement in pain was statistically significant. Clinical important improvements were observed in quality of life, as well as in intensity, duration and frequency of RP. In the experimental group. We believe that this may be related to the sample size and suggest that it is necessary to conduct further studies with larger numbers of patients. This study was carried out in mid-October with the final evaluation conducted in May when the environmental temperature was warmer.

We are convinced that fat micrografts can be enriched with the ADSVF in order to acquire the widely known volumetric effect of lipo-injections as well as the benefits of tissue regeneration produced by the ADSVF. This mixture has already been suggested by Magalon.[25] In our study, the volume infiltrated into each digit was 3 ml. This cellular mixture was administered adjacent to each neurovascular digital pedicle, rather than only into the digital web spaces based on the procedure performed by Bank.[22] Our patients presented with moderately intense vascular and joint conditions and had Cochin scores ranging up to 63. In patients with Cochin scores above 30, the fingers exhibit structured joint stiffness and significant sclerodactyly. Even in these patients, it was possible to administer the cell suspension without causing adverse effects, such as ischemia or compartment syndrome, and without the need for a relaxing incision, as pointed out by Del Papa.[14, 23]

Unlike the conclusion of Malagon,[25] we consider that fat micrografts enriched with the ADSVF are useful for the treatment of mild, moderate, and severe injuries in patients with ischemic hands resulting from SS, including in those with structured joint stiffness. Although no improvements were observed in movement in the present study, additional clinically relevant benefits, such as decreased pain and improved RP, digital ulcer healing were observed.

The site and technique used for acquiring the lipoaspirate affect the quality of the ADSVF in terms of its use for regenerative treatment.[29] Liposuction collection sites have been reported in the knee, hip, femoral region, and abdomen.[14, 24, 25] In our study, fat grafts were mainly obtained from the periumbilical region that is suggested to be the optimal site. Similar techniques have been used for fat acquisition in all studies, including the present work.

Granel et al. processed the ADSVF with a Celution 800/CRS system.[24] Malagon et al. noted that the process requires an evaluation in a cell biology laboratory.[25] In this study, we isolated the ADSVF in a cell biology laboratory. The mean number of viable cells in the ADSVF that we isolated was 228.05 × 10^6^ ± 10.8 compared with 50.5 ± 23.8 (16.7.92.6) in previous studies.[24] Even when there was a larger number of cells in the ADSVF, in addition to those in the fat micrografts, it did not affect our final results.

The techniques performed by the Granel and Magalon group for the characterization and determination of the proportion of the ASCs contained in the ADSVF were consistent with the requirements of the International Federation for Adipose Therapeutics and Science and the International Society for Cellular Therapy and with the Bourin report.[24, 25, 30] In the present study, cell characterization was performed using the following markers: CD34, CD44, CD45, CD73, CD90, CD105, and human leukocyte antigen (HLA)-DR. Their values, together with their adherence to plastic and differentiation toward adipocytes in the laboratory, indicate that the cellular mixture in the ADSVF from our patients, which included mesenchymal stromal cells, endothelial progenitor cells, pericytes, hematopoietic, and immune cells, is similar to those reported previously.[20, 24, 25, 30] Therefore, the obtained results may be explained by the local angiogenic, anti-inflammatory, antifibrotic, immunomodulatory, and regenerative properties of the autologous injected ASCs. It has been demonstrated that the vascular or regenerative properties of the ADSVF are not compromised by SS.[20] Although it is assumed that the administration of the ADSVF may have some systemic effects, the involvement of internal organs did not improve or worsen in association with SS in our patients.

The treated hands of the experimental group patients exhibited a clinically greater volume than the contralateral hands in the same group and hands of the control group patients. This greater volume could prevent the recurrence of ulcers on the bony prominences.

Our study has proven that when combined with conventional medical therapies, the administration of fat micrografts enriched with the ADSVF into the hands of patients with SS can significantly improve pain. Studies with larger numbers of patients are necessary in order to confirm that this method can improve digital ulcer healing, RP, digital perfusion, the vascular density of the nail bed, and the function of the hand.

Based on the aforementioned findings, the regenerative medicinal benefits of the ADSVF are an essential aid for rheumatologists and plastic surgeons before or during the treatment of hand deformities in patients with SS.

## CONCLUSION

The administration of fat micrografts enriched with the ADSVF is a reproducible, safe, and well-tolerated treatment option for the hands of patients with SS. This procedure offers the advantages of the volumetric effect of fat and its regenerative properties. Decreased pain levels and increased healing of digital ulcers, were observed in our patients with SS as well as a tendency toward improvements in RP, and quality of life. Although it is necessary to conduct further studies with larger numbers of patients, we conclude that this treatment is safe and can be administered as a preventive strategy to avoid the progression of hand deformities in patients with SS.

## Supporting information

Supplemental file 3 Statistics

Supplemental File 4 Patient and public involvement

## Data Availability

All data generated or analyzed during this study are included in this published article and its supplementary information files as Additional File 1 Table A, Additional File 2 Table B, Additional file 3 Statistics, Additional File 4 Patient and Public Involvement 1, and Additional File 5 Patient and public involvement 2.

## Abbreviations

SS: Systemic Sclerosis
RP: Raynaud phenomenon
ASCs: Adipose stem cells
ADSVF: Adipose-Derived Stromal Vascular Fraction
DUH: Digital Ulcer Healing
PBS: Phosphate-buffered saline

## DECLARATIONS

### Ethics approval and consent to participate

This research was approved by Institutional Review Committee and The Research Ethical Committee of Instituto Nacional de Ciencias Médicas y Nutrición Salvador Zubirán, with the reference, SCI-1505-15/15-1.

Informed consent to participate in the study was approved by the same committees and was obtained from all the participants.

### Consent for publication

The patients involved in this research in both groups have signed our institutional consent form

### Availability of data and materials

All data generated or analyzed during this study are included in this published article and its supplementary information files as: Additional File 1 Table A, Additional File 2 Table B, Additional File 3 Statistics, and Additional File 4 Patient and Public Involvement 1

## Authors’ contributions

MI: Performed the protocol and the procedure, draft of the manuscript; critically revising the manuscript, data interpretation and discussion of results.

ITV: Isolation of Adipose Derived Stromal Vascular Fraction, and revision of the manuscript.

PBG: Collaboration in the procedure and postoperative control.

TSRR: Performed the protocol and selection of candidate patients.

EATA: Isolation of Adipose Derived Stromal Vascular Fraction, and revision of the manuscript.

ARTP: Performed the protocol and analysis of the results.

AZD: Performed cellular characterization.

MGC: performed the statistical analysis

AMHC: Performed the protocol, enroll patients and obtain informed consent.

## Acknowledgements

The authors thank to Claudia Chavez-Muñoz, MD, for her scientific support, and the Consejo Nacional de Ciencia y Tecnologia (CONACYT) for their funding support.

## Additional files

Additional file 1. Table A. The total viable nucleated cell numbers and cellular characteristics of the ADSVF

Additional file 2. Table B. Complete Results control and experimental groups Additional file 3. Statics. Spreadsheet of data collection

Additional file 4. Patient and public involvement 1

