## SupplementaltableA for "ADIPOSE-DERIVED STROMAL VASCULAR FRACTION AND FAT GRAFTS VERSUS MEDICAL TREATMENT FOR TREATING THE HANDS OF PATIENTS WITH SYSTEMIC SCLEROSIS. A RANDOMIZED CRONTOLLED TRIAL"

| Total viable nucleated cell (x10^6^) | 167.5 (39.8-543) | |
| --- | --- | --- |
| Cell viability (%) | | 82.0 (59.3,95.1) |
| CD34+ (%) | | 4.72 (0.50,18.2) |
| CD45+ (%) | | 43.9 (1.39,67.5) |
| CD44+ (%) | | 36.3(13.8,39.6) |
| CD73+ (%) | | 6.18 (3.91,12.3) |
| CD90+ (%) | | 34.4 (6.43,52.3) |
| CD105+ (%) | | 7.27(1.19,54.9) |
| HLA-DR (%) | | 12.1 (6.26,33.5) |
| Stromal cells (%) | | 4.05 (2.51,6.83) |

Table A. Total viable nucleated cells and characterization of the ADSVF*

*ADSVF: Adipose Derived Stromal Vascular Fraction

Median (min - max)
