## SupplementaltableB for "ADIPOSE-DERIVED STROMAL VASCULAR FRACTION AND FAT GRAFTS VERSUS MEDICAL TREATMENT FOR TREATING THE HANDS OF PATIENTS WITH SYSTEMIC SCLEROSIS. A RANDOMIZED CRONTOLLED TRIAL"

Table B. Complete Results control and experimental groups

| Concept | Day 0 | Day 168 | P^1^ | P^2^ |
| --- | --- | --- | --- | --- |
| **Total Active Motion**  (degrees) |  |  |  |  |
| **Thumb** |  |  |  |  |
| Control | 80(54.2-94.7) | 67.5(54.1-95.8) | 0.72 | 0.68 |
| Experimental | 80(39.4,120) | 77.5(42.5-103) | 0.52 |  |
| **Index finger** |  |  |  |  |
| Control | 160(112,192) | 170(106,185) | 0.15 | 0.32 |
| Experimental | 105 (58.1,178) | 110 (59.8,164) | 0.61 |  |
| **Middle finger** |  |  |  |  |
| Control | 152(116,192) | 170(117,207) | 0.20 | 0.79 |
| Experimental | 130(78.0,198) | 117(78.1,176) | 0.67 |  |
| **Ring finger** |  |  |  |  |
| Control | 177(125,203) | 185(117,207) | 0.81 | 0.76 |
| Experimental | 145 (89.2,198) | 137(86.6,195) | 0.73 |  |
| **Little finger** |  |  |  |  |
| Control | 160(110,218) | 175(109,220) | 0.09 | 0.29 |
| Experimental | 180 (93.2,208) | 127(73.3,185) | 0.20 |  |
| **Digital Sp02 (% oxygen saturation)** |  |  |  |  |
| **Thumb** |  |  |  |  |
| Control | 93.5(91.2,94.7) | 97.0(93.5,97.5) | **0.05** | 0.09 |
| Experimental | 94.0(93.0-96.1) | 95.0 (93.0,96.5) | 0.83 |  |
| **Index finger** |  |  |  |  |
| Control | 94.0(92.0,95.1) | 96.0(93.0,97.8) | 0.07 | 0.30 |
| Experimental | 92.0(85.8,94.7) | 94.5(92.8,95.5) | 0.09 |  |
| **Middle finger** |  |  |  |  |
| Control | 93.0 (88.1,97.1) | 94.0(89.7,96.8) | 0.75 | 0.73 |
| Experimental | 93.5 (89.7-96.2) | 94.5(92.7,96.8) | 0.44 |  |
| **Ring finger** |  |  |  |  |
| Control | 91.5(88.2,93.7) | 94.0(92.2,96.0) | 0.17 | 0.63 |
| Experimental | 91.0(85.5,95.5) | 95.5(93.0,96.5) | **0.05** |  |
| **Little finger** |  |  |  |  |
| Control | 94.0(91.3,95.6) | 92.0 (88.7,95.0) | 0.44 | 0.08 |
| Experimental | 93.5 (87.5,96.3) | 95.0(93.7,96.2) | 0.13 |  |
| **Thumb Opposition** |  |  |  |  |
| Control | 8.50(6.86,8.94) | 9.00(6.93,9.29) | 0.31 | 0.63 |
| Experimental | 7.00(5.16,8.04) | 6.50(4.81,7.99) | 0.58 |  |
| **Pain** |  |  |  |  |
| Control | 4.00(2.64,6.16) | 2.00(1.51,6.04) | 0.91 | **0.02** |
| Experimental | 5.00(3.08,6.12) | 0.00(0.00,2.95)* | **0.006** |  |
| **Raynaud Phenomenon** |  |  |  |  |
| **Frequency, number of event per day/week** |  |  |  |  |
| Control | 5.50(2.68,13.1) | 00(0.00,3.18) | **0.01** | 0.418 |
| Experimental | 4.50(2.87,6.33) | 0.50(0.10,1.10) | **0.005** |  |
| **Raynaud Intensity** |  |  |  |  |
| Control | 2.00(1.22,2.18) | 0.00(0.00,1.44) | **0.03** | 0.60 |
| Experimental | 2.00(1.50,2.10) | 0.50(0.10,1.10) | **0.006** |  |
| **Duration in minutes in every event** |  |  |  |  |
| Control | 17.5(2.12,54.4) | 0.00(0.00,10.3) | **0.02** | 0.14 |
| Experimental | 12.5(6.55,35.6) | 0.00(0.00,6.82) | **0.005** |  |
| **Hand Function (Cochin)** |  |  |  |  |
| Control | 21.0(13.6,39.9) | 23.0(11.8,40.6) | 0.85 | 0.79 |
| Experimental | 21.5 (12.3,38.7) | 23.5(12.5,36.5) | 0.51 |  |
| **Health status and disability index (SHAQ)** |  |  |  |  |
| Control | 0.70(0.53-1.17) | 0.70(0.41-0.95) | 0.40 | 0.32 |
| Experimental | 1.05 (0.61-1.42) | 0.67(0.36-0.90) | 0.11 |  |
| **Quality of life (SF-36)** |  |  |  |  |
| Control | 40.0(28.5,46.4) | 35.0(17.5-51.3) | 0.55 | 0.15 |
| Experimental | 37.5 (32.1-57.8) | 45.0(38.7-62.2) | **0.04** |  |
| **Vascular density of the nail bed** |  |  |  |  |
| Control | 4.62(3.43,5.36) | 4.65(3.70,5.27) | 0.76 | 0.30 |
| Experimental | 4.31(3.46,6.60) | 4.63(3.51,6.26) | 0.33 |  |
| **Skin affection of the hand** |  |  |  |  |
| Control | 6.50(2.32,15.8) | 5.50(1.42,12.9) | **0.05** | 0.19 |
| Experimental | 15.0(6.84,17.7) | 15.5(6.36,16.6) | 0.07 |  |

The sample size of the control group n=9, experimental group n=10; Data are presented as median (95% confidence intervals).

*P<0.05, U de Mann-Whitney analyze the differences between group

P^1^ Analyze the differences between baseline and final was used Wilcoxon signed-rank test.

P^2^	These data were log-transformed before statistical analyses was ANOVA for repeated measures to determine the time x group interaction.
