## Supplemental File 4 Patient and public involvement for "ADIPOSE-DERIVED STROMAL VASCULAR FRACTION AND FAT GRAFTS VERSUS MEDICAL TREATMENT FOR TREATING THE HANDS OF PATIENTS WITH SYSTEMIC SCLEROSIS. A RANDOMIZED CRONTOLLED TRIAL"

*Supplementary file*

*Patient and Public Involvement*

The participation of the patients in this protocol was partially active. They were first contacted by telephone in order for us to acquire information regarding their health status and the evolution of their disease; further, they were asked to express whether they wished to improve any part of their body. In response to questions regarding the function of their hands, the patients stated that if there were a program for improving the function of their hands, they would participate. Based on this information, we explained the objective of the study protocol, to which they agreed. The need for a control group was explained to them. If the results were positive, they would be evaluated by the control patients, and they could make a decision whether or not to undergo the treatment. The interventions, pretreatment, treatment and any discomfort and disabilities that the treatment might cause were fully explained to them. Each of the variables to be evaluated were explained to them. We instructed the patients to evaluate variables, such as coloration, digital oximetry, and pain. They were given a portable digital oximeter, and they recorded oximetry readings every 8 h for the first 3 days, followed 1 day per week. The results were provided to the doctor during the first visit. The evaluation, surgery, and postoperative and total disability durations were explained. Finally, the results were evaluated through validated self-questionnaires. A copy of the document confirming their informed consent and consent to use their data for publication purposes in Spanish is attached.

The patients will participate in recording a standard set of videos that would include the reported patient outcomes to publicize their experiences and share them in intrahospital informative videos at the Instituto Nacional de Ciencias Medicas y Nutricion Salvador Zubiran.
